# Elucidating the Joint Genetic Architecture of Mood Disorder and Schizophrenia

**DOI:** 10.1101/2020.09.14.20193870

**Authors:** Max Lam, Meiling Thompson, Baijia Li, Alexis C. Edwards, Chia-Yen Chen, Tian Ge, Na Cai, Tim Bigdeli, Todd Lencz, Kenneth Kendler, Hailiang Huang

## Abstract

**Introduction:** Recent advances in psychiatric genomics have enabled large-scale genome-wide scans that elucidated genetic architecture both in mood disorder and schizophrenia across individuals of East Asian and European descent. Investigating joint genetic architecture of these psychiatric traits enables the identification of common and diverging etiological mechanisms underlying these psychiatric illnesses. Here, we leverage on the largest GWAS of schizophrenia and mood disorder conducted to date in East Asian and European descent samples to elucidate the joint genetic architecture that underlie these psychiatric disorders.

**Methodology:** We carried out GWAS meta-analysis on both European (EUR) and East Asian (EAS) Ancestry summary statistics for Major Depressive Disorder (MDD) and Schizophrenia via Multi-Trait Analysis of GWAS. Downstream pathway, eQTL, chromatin interaction analysis were carried out to characterize genome-wide results. In addition we carried out genetic correlations and polygenic risk prediction analysis to further study the joint genetic architectures of mood disorder and schizophrenia.

**Results:** There were 308 loci that was significantly associated with at least one trait. Specifically, there were 98 independent loci in EUR-MDD, 5 loci for MTAGx-EAS-MDD, 121 loci for MTAGx-EUR-MDD, 8 independent loci for EAS-SZ, 171 independent loci for EUR-SZ, 124 independent loci for MTAGx-EAS-SZ, and 159 independent loci for MTAGx-EUR-SZ. In all, 61 loci were novel across traits. *SOAT1* and *FOXO3* genes were implicated based on genome-wide associations. 114 gene(s) were implicated in eQTL analysis of gene expression in brain tissue. Gene-set analysis show support for GABA-egic pathways implicated in MDD, driven by several GABA-alpha receptor genes as well as more peripheral *PLCL1* and *NISCH* genes that are responsible for endocytosis and neuronal trafficking. Cross-Ancestry genetic correlations ascertained that the CONVERGE MDD phenotype generally holds higher SNP based heritability and is likely driven by case-ascertainment procedures. Finally, polygenic risk score modelling indicates that MTAGx procedures were effective in enriching GWAS signals in the EAS-MDD for prediction in an independent case-control sample.

**Discussion:** Here we are able to demonstrate that cross-trait cross-ancestry approaches in schizophrenia and MDD not only yields new discoveries to the genetic architecture of these illnesses; we were able to identify new biological underpinnings within the GABA pathways for depressive disorders. The evidence in the current report underscores the importance of taking into consideration both phenotype and ancestry complexities in genome-wide studies.

## Introduction

Major Depressive Disorders (MDD) and Schizophrenia (SZ) are amongst the most debilitating in adult psychiatry; disabilities that are associated with these mental illnesses are associated with significant economic burden (1, 2). There have been hypotheses suggesting that MDD and SZ, tend to be comorbid, with a sizable proportion of individuals with schizophrenia also having diagnosed depression (3, 4). Individuals at-risk to psychosis and first-episode psychosis have been reported to present with affective disorders prior to manifestation of psychosis (5–7). Yet, others have argued that psychosis and/or schizophrenia could be an endpoint of the psychopathology spectrum that begins with significant mood or affective symptom presentation.

Both biological experimentation and genetic studies have demonstrated potential joint genetic architecture that underpins mood disorder and schizophrenia. The biology that jointly underlie SZ and MDD remain poorly understood. Nevertheless, from reviews of earlier genome-wide studies and animal model reports, it appears that MDD and SZ do show evidence of overlap in the area of neurotransmission (dopamine, glutamate, serotonin, gaba-ergic) and neurodevelopmental systems (8–10). In the recent literature large-scale genome-wide association studies have been reported for psychiatric disorders, identifying 102 genomic loci associated with depression (11), 145 loci associated with schizophrenia in European descent individuals [Citation error]; 2 loci associated with depression (12), and 19 loci associated with schizophrenia (13) in individuals of East Asian descent.

Prior reports that investigated the genetic architecture of SZ and MDD indicated a genetic correlation of approximately Rg = 0.45~0.50 within and between European and East Asian ancestry (13). Within MDD, there appears to be differential genetic architecture depending on how cases were being ascertained (14). The phenotype definitions range from MDD that appears recurrent and severe, DSM diagnosed, to depressive symptoms that are self-reported in large-scale biobank collection efforts. Analogous to the phenotype ascertainment is the idea that more severe, recurrent MDD has higher genetic loading. On the other hand, schizophrenia as shown in our previous report tends to be highly consistent in phenotypic ascertainment; as reported, even across ancestries (13). Nevertheless, the significant genetic correlations reported across these traits would suggest that at least to some degree, it would be possible to leverage the overlap of genetic architecture between MDD and SZ despite phenotypic definitions to improve power for discovery.

An added complexity to GWAS meta-analysis over and above phenotypic definitions is the issue of ancestry. In the case of SZ, where cross-ancestry genetic correlation is close to Rg=1, fixed-effect meta-analysis may be utilized to harmonize genetic signals across the genome, without diluting or changing implicitly the inherent phenotype definition. Conversely, when two phenotypes with moderate genetic correlations are meta-analyzed, disentangling if the association signal belongs to one definition or the other; or if by meta-analyzing moderately (genetically) correlated traits have in fact resulted in a “latent” trait makes interpretation challenging. For the case of MDD, which appears to be in the scenario where genome-wide heritability appears differential across studies and ancestry. This is where the Multi-Trait Analysis of GWAS (MTAG; (15)) framework could bring benefits to cross-trait, cross ancestry analysis. The nature of MTAG allows effect sizes to be treated as vectors and projected on an alternate trait based on genetic correlations. This would allow the original trait to be enhanced and overall genetic architecture retained. We describe the MTAG methodology further in the methods section.

In the current report we aim to (1) identify novel genome wide associated loci, that could be enhanced and identified via MTAG (2) carry out genetic correlations that clarifies the phenotype ascertainment in MDD (3) carry out downstream analysis on the results of MTAG and (4) perform polygenic risk prediction in independent case-control samples. The overarching objective is to demonstrate that it is possible to direct GWAS evidence for potential clinical application using a cross-trait, cross ancestry approach even though original discovery GWAS were predominantly conducted in EUR samples.

## Methodology

### Quality control of GWAS summary statistics

Prior to meta-analysis summary statistics we carried out quality control of summary statistics. Prior to quality control procedures there were 9,600,460 variants in the East Asian Mood Disorder ((12), EAS-MDD) GWAS summary statistics; 10,482,735 variants in the East Asian Schizophrenia ((13), EAS-SZ) summary statistics; 8,098,589 variants in the European Mood Disorder ((11), EUR-MDD) summary statistics; and 8,167,164 variants in the European Schizophrenia ((16) EUR-SZ) summary statistics. With the exception of EAS-MDD data, all GWAS summary statistics were obtained from public repositories. To harmonize results of EAS-MDD, which were processed differently from the other results (see (12)), we first converted the raw data into genotypes. Quality control, imputation, and association procedures were then conducted using default parameters via RICOPILI (17). Preliminary results show that the summary statistics obtained via RICOPILI were similar to those in prior reports. For subsequent analysis, we used the version of RICOPILI processed summary statistics, such that data processing procedures were compatible with the other sets of GWAS summary statistics.

Summary statistics quality control is carried out via an in-house quality control pipeline as various sets of GWAS summary statistics were generated over time for separate projects. The summary statistics quality control procedures sought to harmonize genomic position, allele order, strand alignment, allele frequencies across GWAS summary statistics e procedures first aligned CHR, BP coordinates, with A1 and A2 alleles between the 1000 genomes reference panel (phase 3). SNPs with minor allele frequency threshold less than 0.005 were excluded. After allele alignment with the reference panel, variants with allele frequency difference greater than 0.15 were excluded. Finally, we checked strand ambiguous variants against the reference panel. Strand ambiguous variants with allele frequencies differences greater 0.35 were excluded. Finally, variants with allele frequencies that at exactly 0.5 are removed.

### GWAS cross-trait-cross-ancestry meta-analysis

Cross-trait-cross-ancestry analysis was carried out using the MTAG framework. In its original form, MTAG uses LD score regression for estimating the sigma matrix. Along the diagonal of the sigma matrix, are the LD score regression intercept for each trait respectively. While the off-diagonal of the matrix is the bivariate LD score regression intercept. The sigma matrix plays a crucial role in adjusting for potential GWAS inflation due to sample overlap, as well as potential residual population stratification effects (18). The sigma matrix for MTAG can be assembled using the standard LDSC report.

Another crucial aspect of the MTAG framework that estimates bivariate genetic covariances is the general methods of moment approach, which allows MTAG to take advantage of all variant information into account. However, as MTAG is considered a common variant method, genetic covariance could also be estimated using an external genetic correlation approach like LD score regression (19). MTAG allows for external matrices to be estimated and used for estimation of the eventual MTAG estimator albeit such approach is slightly less optimal in that such approaches are more likely to utilize only HAPMAP3 based LD scores rather than all variants present in the input summary statistics. Nevertheless, using externally generated genetic covariance matrices allows a much more flexible framework for MTAG to incorporate various modes of genetic correlations. For purposes of the current study, we investigated the use of the MTAG framework in three stages. First, we demonstrate MTAG-General Methods of Moments (MTAG-GMM) and MTAG-LD Score Regression (MTAG-LDSC) yields overall MTAG Chi-Square values that are largely comparable when combining within ancestry GWAS summary statistics for MDD and SZ. Second, we carry out cross-trait meta-analysis of MDD and SZ within ancestry. The cross trait MTAG yields two sets of GWAS summary statistics (a) MDDsz (b) SZmdd. The trait in capital letters represents the primary trait investigated, while the trait in lower case represents the secondary trait that was projected on the primary trait. The cross-trait analysis was conducted in both EUR and EAS ancestries respectively. Third, we carried out MTAGx, an add-on to the MTAG framework, which estimates cross-ancestry genetic covariance using POPCORN (20). The input summary statistics to MTAGx was (a) MDDszEAS (b) MDDszEUR (c) SZmddEAS (d) SZmddEUR. The output summary statistics are (a) MDDszEASeur (b) MDDszEUReas (c) SZmddEASeur (d) SZmddEUReas. The overall workflow is further visualized in Figure 1. The output summary statistics are defined as follows (a) MDDszEASeur - primary genetic architecture is MDD EAS, secondarily projected on by SZ, and EUR ancestry (b) MDDszEUReas - primary genetic architecture is MDD EUR, secondarily projected on by SZ, and EAS ancestry (c) SZmddEASeur - primary genetic architecture is SZ EAS, secondarily projected on by MDD, and EUR ancestry (d) SZmddEUReas - primary genetic architecture is SZ EUR, secondarily projected on by SZ, and EAS ancestry

**Figure 1.**
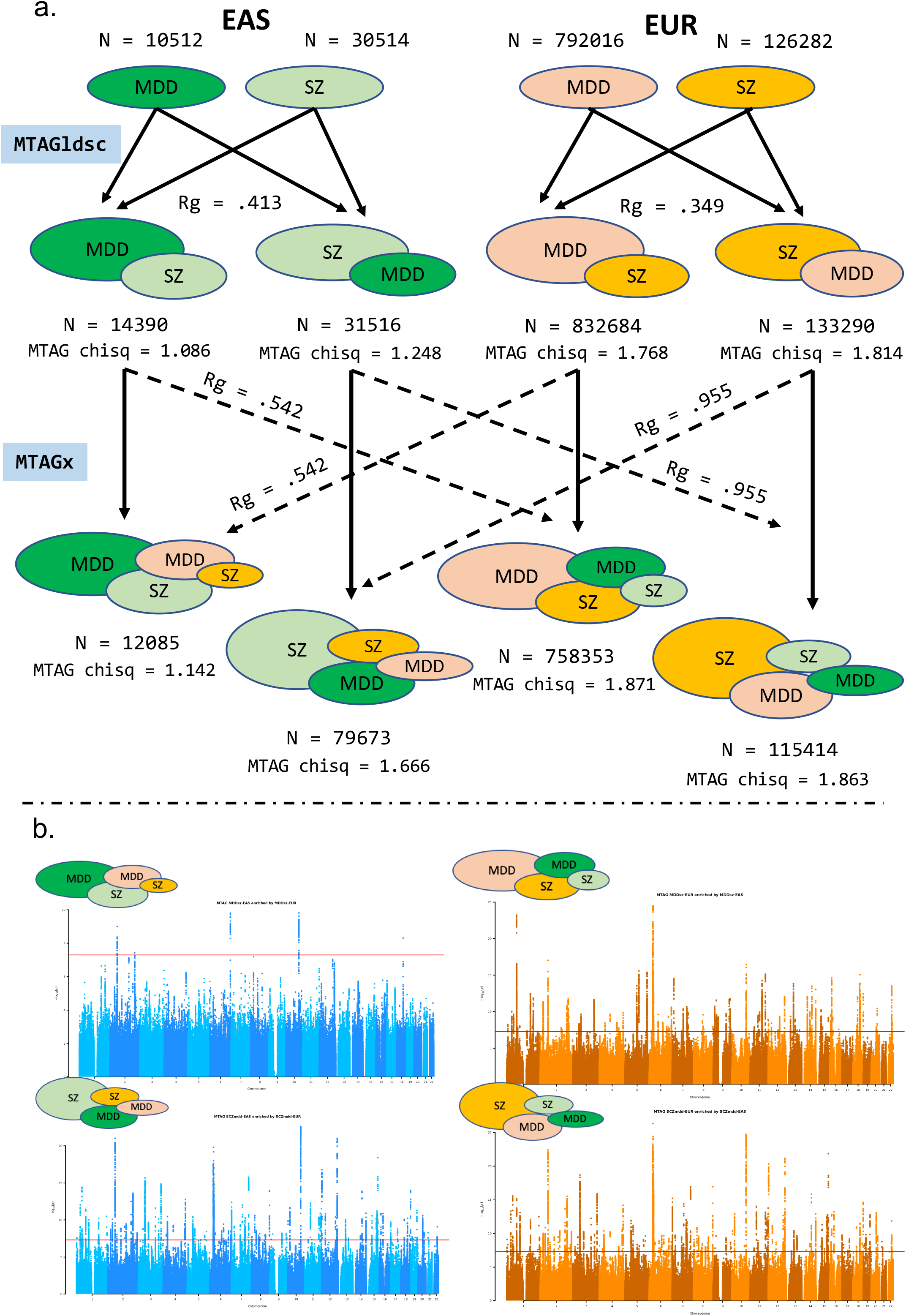
Study design and results for 2-stage enrichment using MTAGldsc and MTAGx. Panel A: MTAGldsc for EAS and EUR ancestry respectively. EAS ancestry MTAG is in two shades of green. Darker green represents EAS-MDD, and the lighter shade of green represents EUR-SZ. Darker shade of orange represents EUR-SZ and lighter shade of orange represents EAS-SZ. MTAGldsc with larger “bubble” represents the primary trait of interest. In the second stage of the analysis, MTAGx was carried out. The largest “bubble” represents the primary trait enriched/projected on by the other traits either from the same ancestry or the alternative ancestry. Panel B: Manhattan plots representing MTAGx outputs. On the top left, MTAGx-EAS-MDD, top right, MTAGx-EUR-MDD. On the bottom left, MTAGx-EAS-SZ and on the bottom right, MTAGx-EUR-SZ.

### Independent SNPs, loci and gene positional mapping

To identify independent GWAS loci joint clumping of all input and output summary statistics were carried out on MDD and SZ GWAS summary statistics for each ancestry respectively, as well as the four MTAG genome wide output indicated in the previous section. GWAS significant threshold of *P* < 5e-8 and LD threshold of R^2^ < 0.1 were used to identify independent significant SNPs. To identify independent genomic loci, SNPs in the LD threshold of R^2^ > 0.6 and with *P <* 0.05 with the independent lead SNPs were extracted. Independent loci were defined based on the lower and upper SNP coordinates of the extracted SNPs. Independent genomic loci that were within 250 kb of each other were then merged into a single locus. LD clumping was carried out using the 1000 Genomes reference phase 3 v 5 reference panel (21) in PLINK 1.90.6. (22). Loci definitions were merged via BedTK (23). Downstream characterization of the GWAS loci was carried out via Functional Mapping and Annotation procedure (FUMA; (24)). Gene positional mapping from the summary statistics were carried out based on ANNOVAR (25) annotations by specifying the maximum distance between SNPs and genes or based on functional consequences of SNPs on genes. The default threshold of 10kb was used. All FUMA output was harmonized using genomic loci definitions via multi-trait joint clumping procedures described above.

### eQTL mapping of Brain Tissue

To identify genes that are jointly expressed in relation to depression and schizophrenia, we leverage on eQTL mapping of gene expression profiled in the various meta-analysis results. These are then consolidated and compared. The following eQTL databases were used for eQTL mapping: (i) GTEx v8 (http://www.gtexportal.org/home/datasets). FUMA contains all SNP-gene pairs of cis-eQTL with nominal P-value < 0.05 (including non-significant associations).

Significant eQTLs are defined as FDR (gene q-value) ≤ 0.05. The gene FDR is pre-calculated by GTEx and every gene-tissue pair has a defined P-value threshold for eQTLs based on permutation. The following tissues were included, Amygdala (N=129), Anterior cingulate cortex (N=147), Caudate (N=194), Cerebellar Hemisphere (N=175), Cerebellum (N=209), Cortex (N=205), Frontal Cortex (N=175), Hippocampus (N=165), Hypothalamus (N=170), Nucleus accumbens (N=202), Putamen (N=170), Spinal cord cervical c-1 (N=126), Substantia nigra (N=114) (ii) PsychENCODE (http://resource.psychencode.org/). The available eQTLs were filtered based on an FDR <0.05 and an expression >0.1 FPKM in at least 10 samples. See Wang et al. 2018 for further details. Ensembl gene ID is used as provided in the original file. The signed statistics are betas (N=1387). (iii) CommonMind Consortium (https://www.synapse.org//#!Synapse:syn5585484). Both eQTLs with and without SVA are included. Available eQTLs from CMC are binned by FDR. Therefore, nominal P-value is not available (replaced with NA). FDR was binned into, <0.2, <0.1, <0.05 and <0.01. Trans eQTLs are also available for CMC data set (as a separated option from cis-eQTLs). Post-mortem brain samples (Frontal Cortex) from 467 Caucasian individuals (209 with SZ, 206 controls and 52 AFF cases (iv) xQTLServer (Dorsolateral prefrontal cortex) (http://mostafavilab.stat.ubc.ca/xqtl/). Gene names were mapped to Ensembl ID (excluded genes which are not mapped to ENSG ID). Since alleles were not available in the original data, extracted from 1000G EUR ancestry based on chromosome coordinate. FDR was not provided in the original data source, but the FDR column was replaced with Bonferroni corrected p-value, as it was used in the original study (corrected for all tested SNP-gene pairs 60,456,556, N=494). (v) Braineac (http://www.braineac.org/). The data included all eQTLs with nominal P-value < 0.05. Since the tested allele was not provided in the original data source, minor alleles in 1000 genome phase 3 are assigned as tested alleles. Signed statistics are t-statistics. eQTLs were identified for each of the following 10 brain regions and based on averaged expression across all of them, Cerebellar cortex, Frontal cortex, Hippocampus, Inferior olivary nucleus (sub-dissected from the medulla), Occipital cortex, Putamen (at the level of the anterior commissure), Substantia nigra, Temporal cortex, Thalamus (at the level of the lateral geniculate nucleus), Intralobular white matter. Expression data was obtained from 134 neuropathologically confirmed control individuals of European descent from the UK Brain Expression Consortium.

### MAGMA and GENE2FUNC gene-set analysis

GENE2FUNC gene set analysis is part of the FUMA pipeline described earlier. To further characterize genes identified by eQTL analysis (described in the previous section), these genes were entered to the GENE2FUNC gene set analysis as exploratory analysis. MAGMA gene association tests and pathway analysis (26) were carried out for each of the MTAG results. 10,678 gene sets (curated gene sets: 4761, GO terms: 5917) from MsigDB v6.2 are used. Benjamini-Hochberg FDR correction was performed for all tested gene sets as part of the MAGMA pathway analysis.

### Within and between ancestry genetic correlation analysis

Genetic correlation analyses were carried out in the quality controlled summary statistics. Cross-ancestry genetic correlations were carried out between EAS-MDD vs EUR-MDD; EAS-SZ vs EUR-SZ via POPCORN ((20)). Within ethnicity genetic correlation was carried out for EUR-MDD vs EUR-SZ; and EAS-MDD vs EAS-SZ via LD-score regression (LDSC;(19)). In either POPCORN or LDSC, SNPs were filtered based on HAPMAP3 variants (~1.2 million SNPs). We utilized default LD-scores computed for the HAPMAP3 SNPs computed on the 1000 genomes phase 3 reference panel ((21)). In addition, we computed liability adjusted heritability scores (h^2) for each of the traits. We also included two additional sets of summary statistics that involved careful curation of mood disorder phenotypes to further investigate how phenotype curation affects cross-trait comparisons of genetic architecture. Additionally, to investigate the effect of phenotypic definitions for MDD, we included GWAS summary statistics for recurrent MDD based on the ICD in the UK Biobank (MDD-UKB-EUR-Recur;(14)) and recurrent MDD based on the DSM-IV in females (MDD-PGC-EUR-Fem-Recur; (27)).

### Polygenic risk prediction of EAS schizophrenia

We carried out polygenic risk score prediction on an independent schizophrenia case-control dataset of EAS ancestry. Patients with schizophrenia and non-psychiatry controls were recruited from multiple institutions (university hospitals and local hospitals) in Japan. The patients were diagnosed according to the Diagnostic and Statistical Manual of Mental Disorders, Fourth Edition (DSM-IV) with consensus from at least 2 experienced psychiatrists. All patients were agreed to participate in the study and provided written informed consent. The study was approved by the institutional review boards of the Tokyo Metropolitan Institute of Medical Science and all affiliated institutions. Genotyping was carried out at The Broad Institute, DNA samples were genotyped on the Illumina Infinium Global Screening Array-24 v1.0 (GSA) BeadChip using standard reagents and HTS workflow procedures procedures for genotyping. GWAS quality control and imputation was carried out via RICOPILI using default parameters. Details of the methodology is reported elsewhere (17). PRS-CS (28), was utilized for polygenic prediction modeling. The method infers posterior effect sizes of SNPs of the four sets of MTAG outputs described earlier. PRS-CS utilizes a high-dimensional Bayesian regression framework, and by placing a continuous shrinkage (CS) prior on SNP effect sizes, robust to varying genetic architectures, provides substantial computational advantages, and enables multivariate modeling of local LD patterns. For the current analysis, we relied on pre-computed LD from the 1000 Genomes Reference Phase 3 reference panel provided with the PRS-CS package. As each set of MTAG summary statistics is projected using both ancestry, which in turn is used for PRS modeling, we carried out shrinkage procedures based on both EAS and EUR ancestry references. Hence, there are two sets of PRS-CS results per prediction modeling analysis. We also carried out PRS-CS modeling where EUR ancestry GWAS summary statistics for MDD and SZ were used for PRS prediction modeling for each of the target databases, representing common practice in the literature where we use as reference points.

## Results

### Quality control and data harmonization overview

We subjected input summary statistics to quality control procedures. After alignment with strand and allele in the 1000 Genomes phase 3 reference panel for the respective ancestries, as well as using quality control filters (See methods); 6,926,896 variants remained for EAS-MDD (N_CASE_ = 5,290; N_CONTROLS_ = 5,223), 7,835,068 variants remained for EUR-MDD (N_CASE_ = 286,534; N_CONTROLS_ = 640,856), 6,494,659 variants remained for EAS-SZ (N_CASE_ = 14,004; N_CONTROLS_ = 16,757) and 7,398,635 variants remained for EUR-SZ (N_CASE_ = 53,386; N_CONTROLS_ = 77,258). These were used as input GWAS summary statistics for MTAG procedures. While we have added on the POPCORN module for genetic covariance estimation within the MTAG framework, standard data harmonization procedures inherent to the MTAG package remain unchanged. As such 5,999,616 variants remained after data harmonization between EAS-MDD and EAS-SZ; 6,891,308 variants remained after data harmonization between EUR-MDD and EUR-SZ. In the second stage, where MTAGx was carried out, 5,034,195 variants remained for the primary mood disorder summary statistics, MTAGx-EAS-MDD and MTAGx-EUR-MDD; and 5,207,548 variants remained for the primary schizophrenia summary statistics, MTAG-EAS-SZ and MTAGx-EUR-MDD.

### MTAGx: Loci Discovery

MTAG allows for bivariate GWAS summary statistics to be projected on each other as vectors. Effectively allowing for GWAS summary statistics for a particular phenotype to retain its genetic architecture, especially if multiple phenotypes are used in MTAG. In the first stage of the analysis, we compared MTAG using the original methods-of-moments procedure for the estimation of genetic covariance using all variants in the summary statistics, against LDSC for the estimation of genetic covariance using HAPMAP3 SNPs. The premise as explained earlier is that most GWAS leverages common variant SNPs for genetic covariance (with pseudo rare variants contributing little to the final Rg). We expected that if sample sizes were large enough, method-of-moments estimation and LDSC will not yield too much of a difference. This was exactly what we found in the case of EUR-MDD and EUR-SZ. MTAG χ^2^ using the methods-of-moments approach was 1.089; with the LDSC approach, MTAG χ^2^ was 1.086. We see the same trend for EAS-MDD and EUR-SZ (MTAG gmm χ^2^ = 1.777; MTAG LDSC χ^2^ = 1.768) (See Figure 1). The preliminary results suggested that it is possible to employ external genetic correlation estimation procedures as extensions to the MTAG framework. As such, we extended the MTAG framework with LDSC and POPCORN, a cross ancestry genetic-correlation method modeled after LDSC. incorporating cross-ancestry genetic covariance estimation allowed cross-trait-cross-ancestry vector projections to be carried out. Figure 1 shows MTAGldsc and MTAGx being carried out in two stages. First, MDD and SZ are projected on each other within ancestry (Rg_EAS-MD_D_-S_z = 0.373; Rg_EUR-M_DD_-S_z = 0.345), Thereafter, we carried out cross-ancestry MTAGx based on cross ancestry genetic correlations (Rg_EAS-EUR-MDD_ = 0.542; Rg_EAS-EUR--SZ_ = 0.955). Post MTAG χ^2^ were as follows: MTAGx-EAS-MDD χ^2^ = 1.142; MTAGx-EUR-MDD χ^2^= 1.871, MTAGx-EAS-SZ χ^2^ = 1.666; MTAGx-EUR-SZ x^2^= 1.863 (See Figure 1). After joint LD clumping, there were 308 loci associated with MDD and SZ across ancestries. Due to the slight differences in clumping parameters and lower number of input SNPs, lead SNPs and loci boundaries differed slightly in the original input GWAS summary statistics. Nevertheless, these differences were small. We categorized loci that were a) not GWAS significant in the input summary statistics of the same phenotype vis-à-vis the primary MTAG phenotype output b) not GWAS significant in any of the input GWAS summary statistics. After harmonization across GWAS significant loci for each GWAS input summary statistics and output MTAG summary statistics, there were 98 independent loci in EUR-MDD, 5 loci for MTAGx-EAS-MDD, 121 loci for MTAGx-EUR-MDD, 8 independent loci for EAS-SZ, 171 independent loci for EUR-SZ, 124 independent loci for MTAG-EAS-SZ, and 159 independent loci for MTAG-EUR-SZ. Of these, there were 40 loci in MTAGx-EUR-MDD, 2 loci in MTAGx-EAS-MDD that were novel to their input summary statistics; while there were 39 loci in MTAG-EAS-SZ and 47 loci in MTAG-EUR-SZ that were novel to their input summary statistics.

In all, 61 loci were novel (Supplementary Table 1, 2). Variants within the novel were ranked via their CADD score > 10, and SNP function. *SOAT1* and *FOXO3*, harboring an exonic variant (rs13306731) and an UTR3 variant (rs111727905) respectively were amongst the top novel signals derived from the MTAG analysis. *SOAT1* was previously associated with Insomnia (29) and *FOXO3* was previously reported to be associated with white matter microstructure in the corpus callosum (30). It should also be note that of the 72 lead SNPs that the 61 loci harbored, 6 of these SNPs (rs9975024, rs337718, rs6983764, rs7811417, rs950394, rs11790388), were also reported in previous EAS schizophrenia studies (13, 31) (Supplementary Table 2). Region plots for the 5 independent significant GWAS loci for MTAGx-EAS-MDD are shown in Supplementary Figures 1-5.

#### Characterization of MTAGx eQTL signals

To further characterize results emanating from MTAGx results we report eQTL signals that are in and around GWAS significant regions (See Supplementary Figure 6). Benjamini-Hochberg FDR correction for multiple testing was carried out on the results. We focus on significantly expressed genes in various brain regions, particularly those that are enriched in EAS-MDD - as MTAGx enables the most significant enrichment for cross-trait-cross-ancestry summary statistics. genes that were unique to MTAGx-EAS-MDD as these would represent gene-sets associated with the more severe MDD phenotype in the EAS ancestry. 114 genes were significantly expressed in brain tissue, that underlie the MTAGx-EAS-MDD results. Importantly, eQTL analyses support earlier variant based annotations of *SOAT1* and *FOXO3*. Further examining the 114 genes, 54 (47%) of them overlapped the MTAGx-EUR-MDD results, 45 (49%) overlapped uniquely with schizophrenia results (from either ancestry) (See Figure 2). It is notable that several of these genes *AS3MT, CNNM2, NT5C2, ZNF804A*, are also known to be associated with bipolar disorders and mood disorder reported in earlier cross-disorder analysis (32, 33). To further understand the biological underpinnings of the expressed genes, hypergeometric tests were conducted of the 114 genes against protein coding genes. We selected only GO-gene-sets with the Molecular Signature database 6.2 to facilitate comparisons with subsequent MAGMA gene set analysis. The preliminary gene-set analysis here suggests that crucial neuronal regulatory pathways were implicated (See Figure 2). Strikingly, the 5 significant associations that underlie the results of MTAGx-EAS-MDD (Supplementary Figures 1-5) show not only GWAS significant associations primarily related to the EAS-MDD but also appear to be in highly functional regions underscored by the existence of large swathes of eQTL signals and HiC annotations.

**Figure 2.**
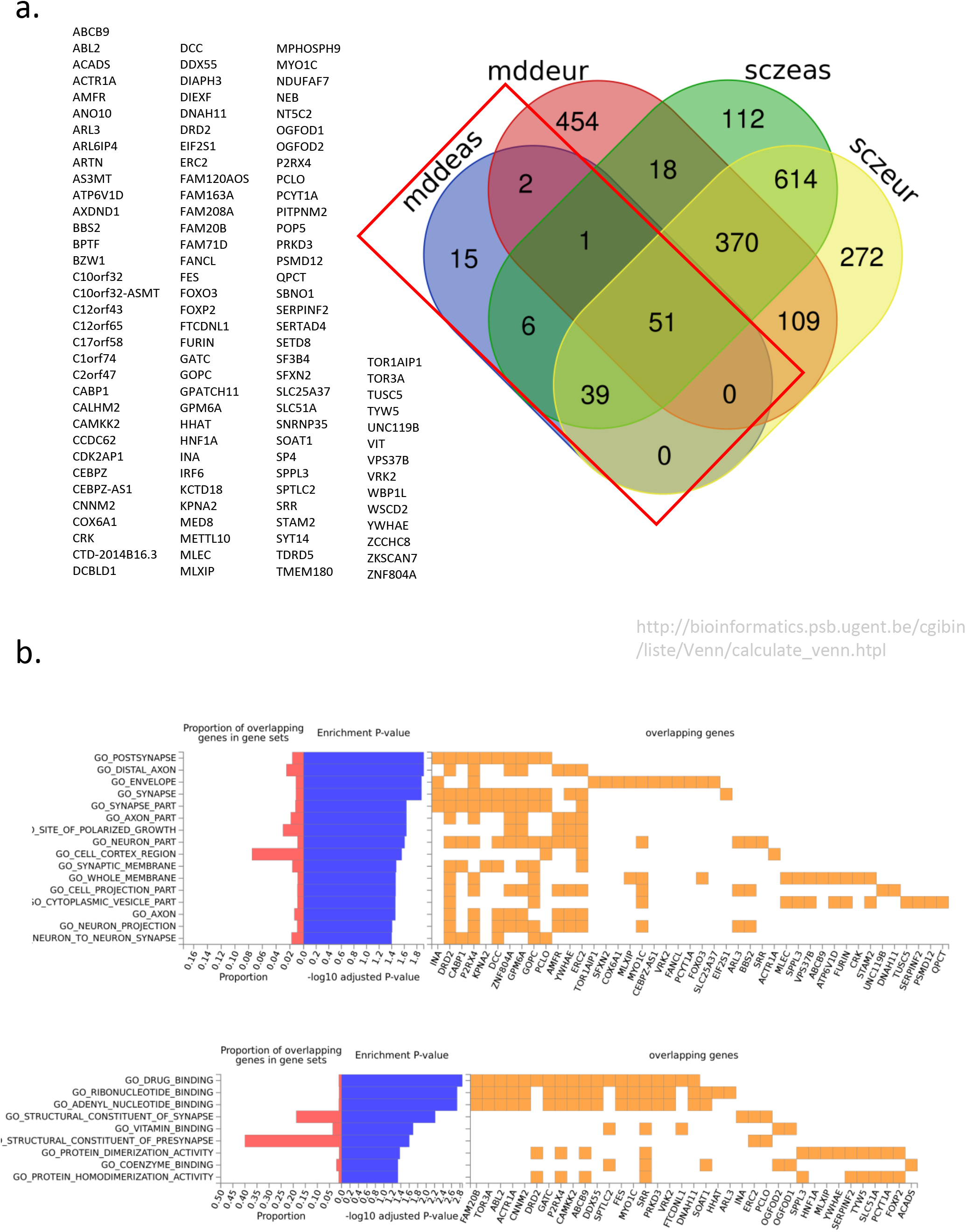
eQTL significant genes for MTAGx results and Hypergeometric Gene-Set analysis. Panel A: 114 significant expressed genes in brain tissue represented in MTAGx-EAS-MDD. Venn diagram shows MTAGx output for EAS-MDD, EUR-MDD, EAS-SZ, and EUR-SZ. Panel B: Hypergeometric gene-set analysis for 114 gene against GO gene ontologies for Biological, Cellular, and Molecular functions.

#### MAGMA gene set analysis

MAGMA gene set analysis was performed for each of the MTAGx results. Gene-set analysis was conducted based on the gene mapping Gene-set analysis was conducted based on gene mapping procedures on the MTAG output summary statistics. For each, the Molecular Signature Database (v6.2) was used as the annotations. Results were then combined across the MTAG output (Supplementary Table 4). Gene set analyses indicate several biological systems being implicated, particularly synaptic, and neuronal pathways. Some of these have been previously reported in mood disorder and schizophrenia literature. Nonetheless, the GABA-ergic pathway emerging for the MTAGx-EUR-MDD results (MAGMA_Z_ = 4.32; MAGMA_P-FDR_ = 0.0021), appears to be in support of GABA dysfunction in mood disorders. We further examined genes that were part of the GABA-gene set to identify genes that are implicated in the pathway, and also searched eQTL results that might also be implicated as part of the GABA-signalling. The following genes have been implicated *DRD2, GABRA4*, *GABRA6, GABRB2*, *GABRG3, NISCH, PLCL1*, *PLCL2* (See Figure 3, Supplementary Table 3, 4).

**Figure 3.**
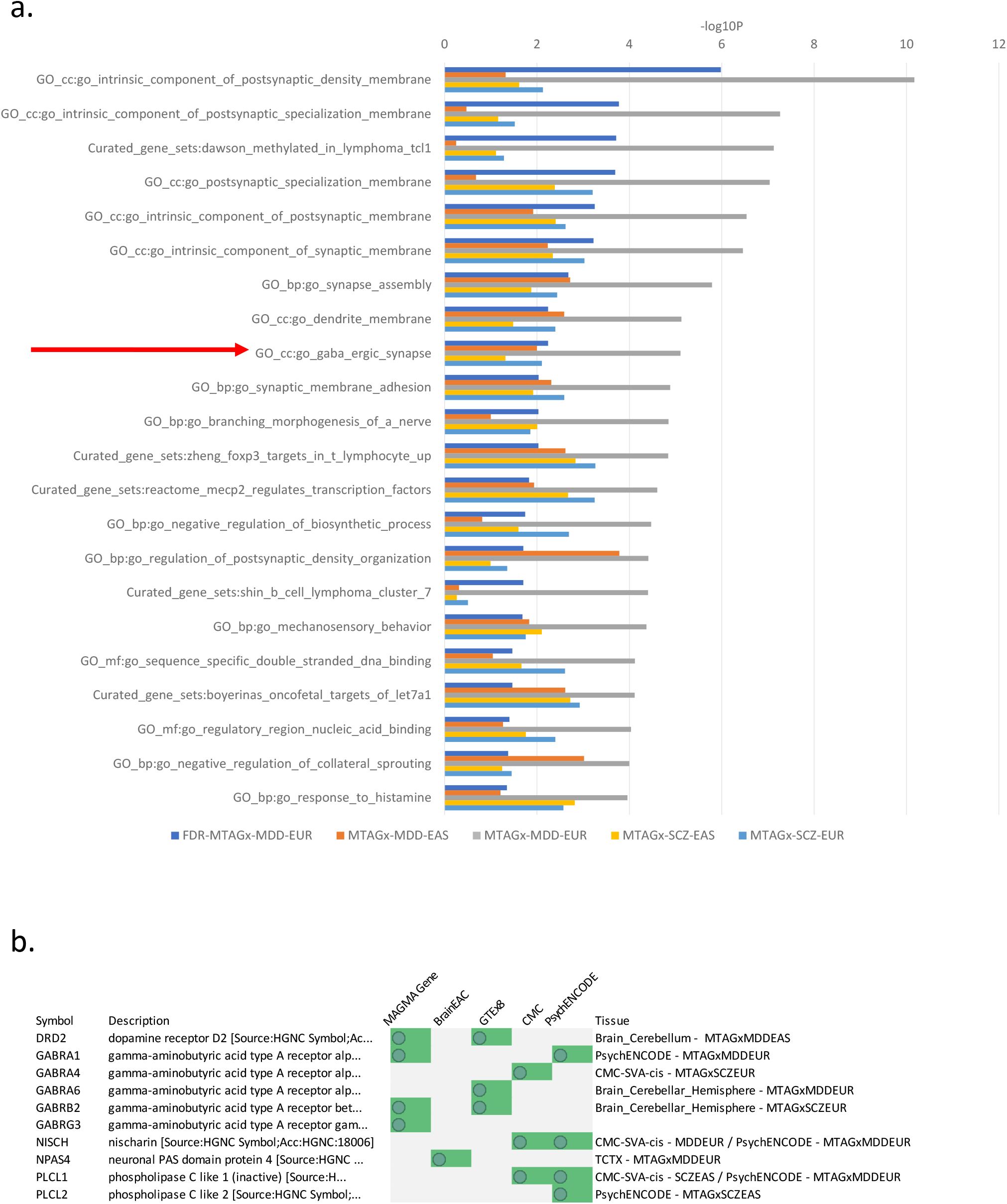
MAGMA gene-set analysis for MTAGx results. Panel A: Significant MAGMA gene-set results. In darker blue, is the MTAGx-MDD-EAS results after Benjamini-Hochberg multiple testing correction. Other colored bars represent gene-set analysis results from each of the MTAGx outputs. Panel B. GABA-ergic synapse/signalling gene-set with supporting gene-based results either from gene-mapping of MTAGx or eQTL results. *Note:* Error bars represent 95% Confidence interval.

#### Genetic correlation

In the earlier section, we leverage cross ancestry correlations carried out within the POPCORN package with the MTAG framework to allow for cross-trait-cross-ancestry enrichment of GWAS association signals. In this section, we carry out POPCORN to investigate the SNP based heritability, as well as genetic correlations of the EAS-MDD data set with other similarly curated phenotypes reported in previous studies (11–14, 27). Genetic correlation analyses show that the observed heritability for schizophrenia in both ancestries are expectedly high (Rg_EUR-SZ_ = 0.70; Rg_EAS-SZ_ = 0.69). Our expectation that EAS-MDD being a more clinically driven phenotype is supported in that the heritability appears significantly higher than MDD phenotypes derived in large scale biobank studies, i.e. Rg_EAS-MDD_ = 0.48 is comparable to females clinically diagnosed with MDD via DSM-criteria rather than minimally phenotyped MDD (See Figure 4A). Moreover, though variant heritability matches recurrent MDD in females reported in a previous PGC cohort, we note that genetic correlation between these phenotypes continue to be modest compared to schizophrenia. These genetic correlation results further supported the rationale for an MTAGx analysis rather than standard inverse variance GWAS meta-analysis. The results also would inform subsequent polygenic risk prediction analysis reported in the subsequent section.

**Figure 4.**
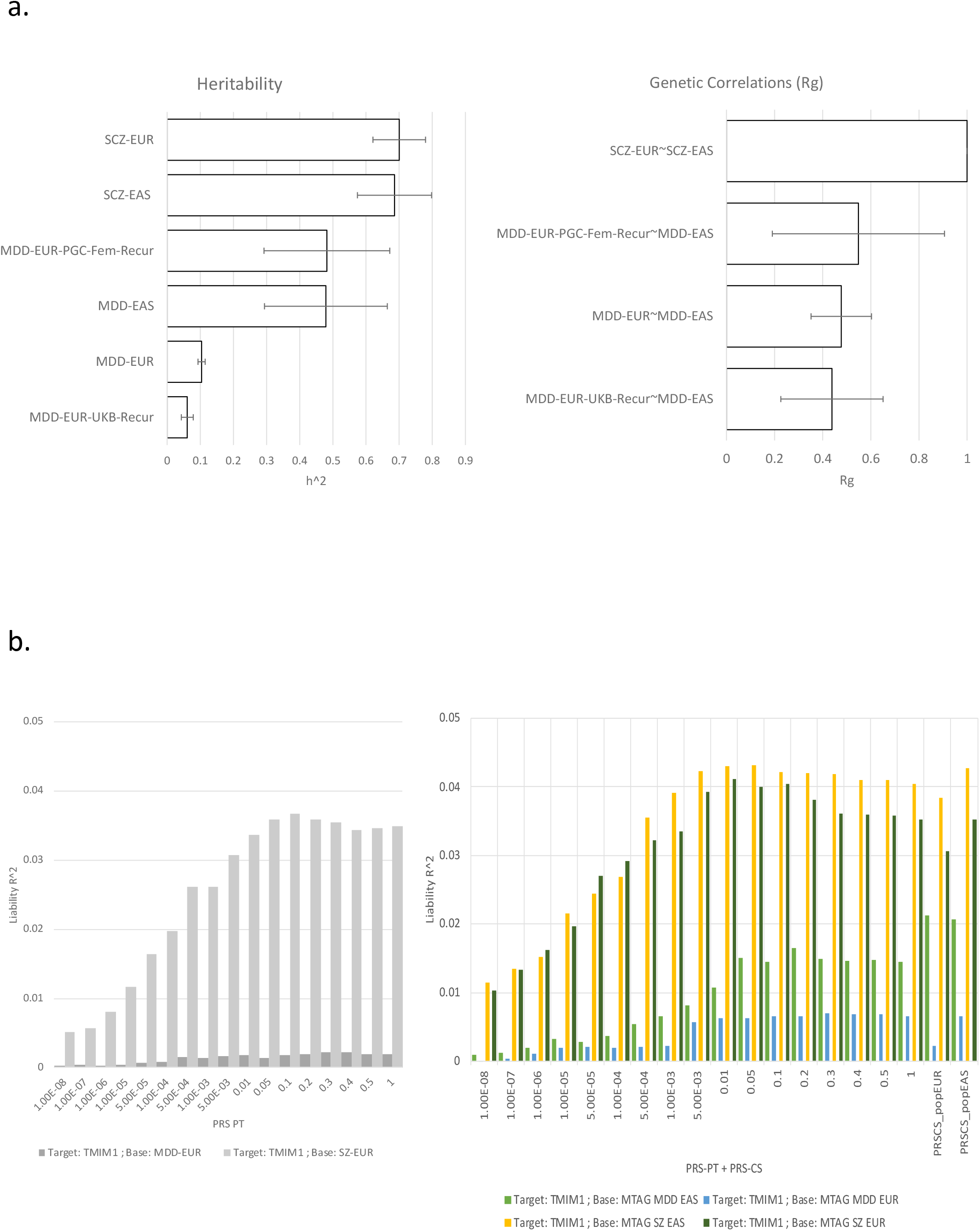
Transethnic Genetic Correlations and Polygenic Risk Score modelling. Panel A: Left-hand panel, are the SNP heritability for each trait. Right-hand panel, genetic correlations obtained via POPCORN. Panel B: Left-hand panel, standard P_T_ genetic risk score modelling using EUR-MDD and EUR-SCZ as base data for prediction into independent GWAS EAS schizophrenia case-control cohort. Right-hand panel, MTAGx results used as base data for genetic risk prediction into the same GWAS EAS schizophrenia case-control cohort.

#### Polygenic Risk Modelling

Polygenic risk modelling was carried out to predict schizophrenia case-control status based on each MTAGx output. We first carried out PT analysis using both EUR-MDD and EUR-SZ summary statistics as base data for predicting into a target schizophrenia case-control dataset (See Figure 4B). PT results show an expected trend, with the polygenic signal saturating with more SNPs. Nevertheless, EUR-MDD though significant does not appear to be optimal as a predictor, with effect sizes much smaller than schizophrenia, which is expected. The intuition for the MTAGx output as base predictors into the same schizophrenia case-control data is that by enhancing GWAS associations representing the trait of interest, we would expect improvement in polygenic risk score prediction. This was exactly the case. We found that the MTAGx-EUR-MDD based statistics improved by 2.19 times in its prediction of case-control status, while MTAGx-EAS-MDD showed the most dramatic improvement in polygenic risk prediction by 6.59 times. Both MTAGx EAS and EUR SZ improved by 1.17 and 1.12 times compared to EUR-SZ only prediction which is not unexpected. In addition, we carried polygenic risk prediction modelling using PRS-CS. The results of PRS-CS largely agreed with results of PT polygenic risk modelling. However, it is notable that in some cases PRS-CS showed better performance for PRS prediction when the reference panel is tuned to the target sample rather than the MTAGx output.

## Discussion

Previous reports of genome-wide association for MDD have yielded evidence that mood disorder like schizophrenia is highly polygenic, and implicates brain and neuronal level biological underpinnings. More recently, evidence has also emerged that unlike schizophrenia, where observed SNP heritability is sufficiently high, and that phenotype ascertainment is relatively uniformed; phenotyping in MDD tends to have an influence on the observed genetic effects. It is apparent, but not surprising, that more severe recurrent MDD comprises higher genetic loading compared to less rigorous phenotyping procedures e.g. self-reports existent in large-scale biobanks. Amidst calls for greater diversity and inclusion of other ancestries within studies of psychiatric genomics, our group has initiated the largest cross-ancestry research efforts in the Han Chinese ancestry for schizophrenia. Nevertheless, there continues to be opportunities to expand efforts for MDD. The CONVERGE data, previously presented, yields a unique opportunity to study the effects of phenotype ascertainment and cross-ancestry work in genomics in tandem.

As prior reports have discussed, phenotype ascertainment in the CONVERGE data had been significantly more rigorous for MDD than other sites that have reported large-scale GWAS evidence. Arguably, phenotyping procedures in the CONVERGE data could be thought of as “optimizing” the sample for genetic discovery given the sample inclusion criteria for recurrent mood disorders, and in predominantly females. This appears very evident in the genetic correlations and heritability comparisons reported in the earlier sections. Though with modest sample sizes, we observe that the CONVERGE (i.e. EAS-MDD) show a much higher mean SNP heritability as compared to a large-scale GWAS of MDD close to a million individuals. Moreover, when compared to a more carefully ascertained sample of EUR ancestry, matching that of the CONVERGE data, we observe similar SNP heritability; further supporting the notion that the phenotype reported with the CONVERGE dataset is likely to be much more genetically loaded clinical phenotype. As such, we sought to further leverage existing cross-ancestry data for schizophrenia in an attempt to enrich GWAS association signals within the CONVERGE data using an extension of MTAG. It should be noted that we selected MTAG as a framework for cross-ancestry-cross-trait enrichment of GWAS association signals because MTAG tends to preserve the existing genetic architecture of the input data. This is compared to other methods such as the Hans and Eskin (xx) Random-Effects approach (34), or more recently PLEIO (35) that uses a similar approach but combines data similar to GWAS meta-analysis. In the case of the latter two approaches, cross-trait enrichment becomes challenging, as the implicit assumption of these cross-trait methods is elucidating pleiotropy, but combining cross-trait GWAS summary statistics inadvertently results in a “latent-trait” that is either equally weighted, or driven by the trait with larger samples/statistical power.

By “borrowing” statistical power, we were able to demonstrate that MTAGx is an effective tool for loci discovery. Of the 308 loci elucidated from the analysis 61 of them were not reported in the original input summary statistics. 6 of the 72 SNPs harbored within the 61 loci, were reported in large-scale cross-ancestry GWAS of schizophrenia. This indicates that MTAGx methodology not only captures what tends to be reported via transethnic meta-analysis, it facilitates loci discovery to a greater extent leveraging precisely on the cross-trait-cross ancestry approach. The primary objective of enhancing the genetic architecture of EAS-MDD from the CONVERGE data, was met. Importantly, 5 loci were identified and of those 5, two, were novel to MDD. We also acknowledge however that some of the loci were also shared with schizophrenia. What appears noteworthy is at least one of the loci within the MTAGx-EAS-MDD significant loci is a known Bipolar disorder genomic region (i.e. *CNNM2, AS3MT)*. While further work is necessary to further understand each of the 5 regions identified, it is possible that enhancing the genetic architecture of more rigorously defined mood disorder phenotype, via MTAGx with schizophrenia, and more generalized mood disorder phenotype, we were able to index loci that tends have greater biological sequelae.

We further attempted to understand the biological underpinnings of genes identified in brain expressed eQTL as well as genes that are mapped more broadly from the MTAGx results. We identified several neuronal regulation pathways based on genes with significant eQTL signals in MTAGx-EAS-MDD. These included (list pathways). The results are largely consistent with what is observed with the MAGMA gene set analysis. Although, it is necessary to point out that MTAGx-EAS-MDD was not powered enough for MAGMA gene-set analysis. Results from the gene-set analysis are mainly driven by the other MTAGx results. The gene-sets that emerged as significant for MTAGx-EUR-SZ, MTAGx-SZ-EAS and MTAGx-SZ-EUR were largely consistent (list pathways). Worth highlighting are the results from MTAGx-EUR-MDD where we observed that GABA-ergic signalling emerged as a significant gene-set. This is not trivial given that previous studies did not find this gene-set to be significant. It is likely that the GABA-ergic signaling pathway is emerging due to the increased power from the EAS and schizophrenia datasets included in the MTAGx analysis. Though not surviving multiple corrections in the MTAGx-EAS-MDD gene-set analysis, we note that it shows a trend result. The results here support GABA dysfunction hypothesis in mood disorders. It could also be likely that the GABA signalling gene-set is elucidated due to enrichment from the more clinically severe phenotype ascertained as part of the CONVERGE data. Examining genes that are driving the gene-set analysis results we found that genes coding for GABA-alpha subunit 1, GABA-beta subunit 2 and GABA-gamma subunit 3 were significant. These subunits are part of GABA-A-receptors that tend to be activated by chloride ion, regulating GABA signalling across neurons. Other eQTL results for MTAGx-EUR-MDD also implicated genes encoding the Phospholipase C-related but catalytically inactive protein and nonadrenergic imidazoline-1 receptor protein. Both genes *PLCL1* and *NISCH* are part of the GABA-ergic signalling pathway but play more peripheral regulatory roles such as membrane trafficking, and endocytosis for GABA. It is, however, necessary in subsequent work to establish the exact variant that is directly causal to these genes, its downstream effects on the pathways.

Lastly, we investigated polygenic prediction into an independent schizophrenia dataset. First, we carried out polygenic prediction modelling using the P-thresholding approach based on the large GWASs of EUR-MDD and EUR-SZ, to establish a baseline that we could compare against. This represented the standard polygenic risk prediction analysis that is expected in most research contexts. Next, we used each of the MTAGx outputs as base data for prediction. A crucial aspect is that sample size is not considered an issue. The reason was that MTAGx did not significantly increase the sample sizes of the traits that we were most interested in MTAGx-EAS-MDD and MTAGx-EUR-MDD. We expected the MTAGx schizophrenia base to perform well, based on the baseline PT modelling results, and that was supported by the subsequent MTAGx results - however, this did not seem surprising, given that the target data is in fact a schizophrenia case-control dataset. However, there was a significant improvement when we applied the MTAGx MDD datasets as the base predictors. Despite being less powered based on estimated sample sizes in the MTAGx-EUR-MDD results, there was an improvement in polygenic prediction. The improvement in prediction is much more pronounced with the MTAGx-EAS-MDD which is potentially driven by the more rigorous phenotyping in the CONVERGE data. We also carried out PRS-CS which purports to improve prediction accuracy via Bayesian methods. What appears to be notable from PRS-CS results is that for MTAGx results, it is more preferable for PRS-CS to be tuned according to the ancestry of the target data. Further work is necessary to clarify this curious phenomenon.

## Concluding Remarks

The results reported in here represents the first data freeze of ongoing analysis that the authors wish to share with the community. Preliminary findings suggest that MTAGx is effective in enhancing the original genetic architecture of input data. This allows improvement in loci discovery, which improves downstream eQTL and gene set analysis. Importantly, MTAGx serves to improve polygenic risk prediction that allows larger, more powered EUR based GWAS to be used for enhancing smaller more modest EAS results. Additionally, it would also allow for traits that have higher SNP heritability to be exploited for polygenic risk prediction analysis.

## Data Availability

This is the first data-freeze of the manuscript. GWAS summary statistics reported in the pre-submission version of the manuscript will be provided.

## IRB Statement of Approval

We have used open sourced/de-identified GWAS summary statistics for the main data analysis reported in the current study. For those no IRB oversight is necessary. For the Asian GWAS genotype data that was used for the polygenic risk prediction, the IRB statement is as follows:. All patients were agreed to participate in the study and provided written informed consent. The study was approved by the institutional review boards of the Tokyo Metropolitan Institute of Medical Science and all affiliated institutions.

## Acknowledgements

Funding Statement:

HH acknowledges support from the Zhengxu and Ying He Foundation and the Stanley Center for Psychiatric Research.

## Supplementary Materials

Supplementary Tables

Supplementary Table 1: Significant Genomic Loci for each dataset, MDD-eas-input; MDD-eur-input, SCZ-eas-input; SCZ-eur-inpu; MTAG-MDD-EAS-output; MTAG-MDD-EUR-output; MTAG-SCZ-EAS-output; MTAG-SCZ-EUR-output

Supplementary Table 2: Independent significant GWAS loci for each dataset, MDD-eas-input; MDD-eur-input, SCZ-eas-input; SCZ-eur-inpu; MTAG-MDD-EAS-output; MTAG-MDD-EUR-output; MTAG-SCZ-EAS-output; MTAG-SCZ-EUR-output. *Note:* Novel Loci reported in second tab, VEP annotations reported in third tab.

Supplementary Table 3: eQTL gene expression information for MTAGx output for mddeas, mddeur, sczeas and sczeur. BrainEAC, GTEx8, CMC, PsychEncode, eQTLCatalogue, xQTL are reported in this supplementary table.

Supplementary Table 4: MAGMA gene based analysis for MTAG summary statistics. MAGMA gene set analysis results for MTAG-MDD-EAS, MTAG-MDD-EUR, MTAG-SCZ-EAS, MTAG-SCZ-EUR.

Supplementary Figures

Supplementary Figures 1-5: Region plots for GWAS significant signals for MTAGx-EAS-MDD.

Supplementary Figure 6: Mixed manhattan plot for eQTL signals. Panel A: BrainEAC; Panel B: GTEx8; Panel C: PsychENCODE.

